# A genetic exploration of the relationship between Posttraumatic Stress Disorder and cardiovascular diseases

**DOI:** 10.1101/2024.03.20.24304533

**Authors:** Eva Lukas, Rada R Veeneman, Dirk JA Smit, CHARGE Inflammation Working Group, Jentien M Vermeulen, Gita A Pathak, Renato Polimanti, Karin JH Verweij, Jorien L Treur

## Abstract

**Background and Aims:** Experiencing a traumatic event may lead to Posttraumatic Stress Disorder (PTSD), including symptoms such as flashbacks and hyperarousal. Individuals suffering from PTSD are at increased risk of cardiovascular disease (CVD), but it is unclear why. This study assesses shared genetic liability and potential causal pathways between PTSD and CVD.

**Methods:** We leveraged summary-level data of genome-wide association studies (PTSD: N= 1,222,882; atrial fibrillation (AF): N=482,409; coronary artery disease (CAD): N=1,165,690; hypertension: N=458,554; heart failure (HF): N=977,323). First, we estimated genetic correlations and utilized genomic structural equation modeling to identify a common genetic factor for PTSD and CVD. Next, we assessed biological, behavioural, and psychosocial factors as potential mediators. Finally, we employed multivariable Mendelian randomization to examine causal pathways between PTSD and CVD, incorporating the same potential mediators.

**Results:** Significant genetic correlations were found between PTSD and CAD, HT, and HF (*r_g_* =0.21-0.32, p*≤* 3.08 *·* 10*^−16^), but not between PTSD and AF. Insomnia, smoking, alcohol dependence, waist-to-hip ratio, and inflammation (IL6, C-reactive protein) partly mediated these associations. Mendelian randomization indicated that PTSD causally increases CAD (IVW OR=1.53, 95% CIs=1.19-1.96, p=0.001), HF (OR=1.44, CIs=1.08-1.92, p=0.012), and to a lesser degree hypertension (OR=1.25, CIs=1.05-1.49, p=0.012). While insomnia, smoking, alcohol, and inflammation were important mediators, independent causal effects also remained.

**Conclusions:** In addition to shared genetic liability between PTSD and CVD, we present strong evidence for causal effects of PTSD on CVD. Crucially, we implicate specific lifestyle and biological mediators (insomnia, substance use, inflammation) which has important implications for interventions to prevent CVD in PTSD patients.

**Translational perspective:** The significant mental and physical strain experienced by patients suffering from Post-traumatic Stress Disorder (PTSD) remains a domain necessitating further insight for the development of effective intervention strategies. Our study elucidates the complex genetic architecture that underlies the relationship between PTSD and cardiovascular disease. We present evidence supporting a causal link from PTSD to coronary artery disease and heart failure. Further, we identify various mediators of this causality, including inflammatory markers, substance use, waist-to-hip ratio and sleep deprivation. Our work calls for targeted preventive and therapeutic approaches to reduce the dual burden of mental and physical disease in PTSD patients.

## Introduction

Posttraumatic stress disorder (PTSD) is a disruptive mental illness triggered by traumatic experiences and manifests as recurrent, adverse mental states related to the event. Symptoms may include flashback nightmares, dissociation and/or physiological reactions to external or internal event-related triggers. Traumas that may cause PTSD include acts or threats of (sexual) violence and witnessing the man-r disaster-caused passing of others [1]. While PTSD can only develop if a traumatic event has been experienced, the risk of developing it is partly determined by genetic liability. Twin-based studies estimated PTSD heritability between 40-60% and genome-wide association studies (GWAS) demonstrated that PTSD is characterized by high polygenicity, signifying that the risk of developing the disorder involves the influence of numerous genes [2, 3].

On top of debilitating psychological symptoms, PTSD has been associated with an increased risk of cardiovascular disease (CVD), as reported in prior observational research [4, 5]. Multiple studies and reviews have underscored an increased prevalence of diverse cardiovascular conditions among individuals with PTSD, including ischemic heart disease, heart failure, and atrial fibrillation [6, 7]. Why PTSD is associated with CVD is not fully understood. One possibility is that the development of PTSD and the development of CVD are (partly) due to the same genetic risk factors [8]. It could also be the case that there are causal associations, with PTSD acting as a risk factor for CVD, potentially mediated by biological (e.g. hypothalamic-pituitary-adrenal dysregulation, lower cortisol levels in the period following trauma, higher heart rates, elevated blood pressure, increased inflammation), behavioral (e.g. substance abuse, obesity, sleep disturbance) and/or psychosocial (e.g. social isolation) factors (extensively reviewed in [9] and [10]). Conversely, it may also be possible that a diagnosis or experience of life-threatening CVD could provoke PTSD [4]. It should be noted that while the DSM-4 considered life-threatening illnesses as PTSD triggers, the DSM-5 has limited these to unexpected medical catastrophes, like waking during surgery [1, 11].

We used advanced genetic methods to investigate the nature of the association between PTSD and CVD, focusing on shared genetics and causal explanations. In this study, we had two primary objectives: Firstly, we explored the shared genetics between the two conditions. Secondly, we established the directionality of a potential causal link and subsequently elucidated underlying mechanisms, mediating causal relationships. To investigate this, we leveraged data from existing large-scale GWAS. These studies assess the association of millions of single-base DNA changes called single-nucleotide polymorphisms (SNPs) with a certain trait [12]. By integrating GWAS data from multiple traits, one can unveil genetic associations and potentially establish causality between different phenotypes [13].

## Methods

The analysis plan was preregistered (for updates and alterations of the preregistered datasets please see Supplementary notes). We exploited summary-level data of large GWASs of European ancestry and applied advanced genetic methods to unravel the complex relationship between PTSD and CVD. PTSD genetic information was derived from the GWAS meta-analyses from the Psychiatric Genomics Consortium [3]. For the CVD traits, we selected atrial fibrillation (AF) [14], coronary artery disease (CAD) [15], hypertension (HT) [16] and heart failure (HF) [17] (Table 1).

**Table 1:** Summary-level GWAS data used for analysis. Effective sample size noted in brackets where available. UK-Biobank abbreviated as UKB.

| Phenotype | Measure | Study | Sample Size (Thousands) | SNPs |
| --- | --- | --- | --- | --- |
| PTSD | clinician administered, self-report, DSM-4 Posttraumatic Stress Disorder Checklist, electronic health records | Nievergelt et al., (2023) [3] | 1,222.9 (1,085.7) | 73 |
| Atrial Fibrillation | Paroxysmal or permanent AF, atrial flutter, | Roselli et al., (2018)[14] | 537.4 (482.3) | 84 |
| Coronary Artery Disease | See original Supplementary Table 32 | Aragam et al., (2022) [15] | 1,165.7 (984.2) | 241 |
| Hypertension | UKB | Zhu et al., (2019) [16] | 458.6 (313.8) | 22 |
| Heart Failure | Clinical diagnosis, any etiology | Shah et al., (2020) [17] | 977.3 (930.0) | 12 |
| **Biological factors** |  |  |  |  |
| Serotonin | Plasma levels | Hysi et al., (2022) [20] | 8.8 |  |
| Cortisol | Morning plasma cortisol levels | Crawford et al., (2021) [21] | 25.3 | 4 |
| Growth Hormone Receptor | Plasma protein level | Sun et al., (2018) [22] | 3.3 | 1 |
| BDNF | Serum and plasma levels | Li et al., (2020) [27] | 11.8 | 7 |
| C-reactive Protein | UKB | Said et al., (2022) [23] | 575.5 | 266 |
| IL-6 | Blood samples | Ahluwalia et al., (2021) [24] | 52.7 | 9 |
| IL-8 | Serum or plasma levels | Folkersen et al., (2020) [25] | 21.8 | 1 |
| TNF-α | | Sliz et al., (2019) [26] | 13.6 | 1 |
| **Behavioural factors** |  |  |  |  |
| Alcohol Intake | Drinks per week | Saunders et al., (2022) [28] | 2,428.9 | 496 |
| Alcohol Dependence | DSM-4 diagnosis | Walters et al., (2018) [29] | 46.6 (35.0) | 2 |
| Smoking Initiation | Ever smoked regularly | Saunders et al., (2022) [28] | 2,669.0 | 1,346 |
| BMI | UKB | Yengo et al., (2018) [30] | 700 | 941 |
| waist-hip-ratio | UKB | Pulit et al., (2019) [31] | 694.6 | 346 |
| Insomnia | UKB | Watanabe et al., (2019) [32] | 386.1 | 31 |
| **Psychosocial factors** |  |  |  |  |
| Loneliness | UKB | Abdellaoui et al., (2019) [33] | 511.3 (430.3) | 16 |
| Educational Attainment | years-of-education, UKB | Okbay et al., (2022) [34] | 3,000.0 | 131 |
| Educational Attainment | years-of-education | Okbay et al., (2016) [38] | 293.7 | 74 |

### Genetic correlation and latent factor analysis

We applied Linkage-Disequilibrium Score Regression (LDSC) [18] in a genomic structural equation modeling (Genomic SEM) framework [19] to assess genetic correlation and common variance between PTSD and CVD. In essence, LDSC quantifies the extent to which genetic variants are linked to phenotypic traits. LD scores represent the correlation between a genetic variant and nearby variants. LDSC calculates shared genetic influences between traits estimating genetic correlation to be negative (opposing effects), 0 (no overlap) or positive (overlap). The slope in LDSC regression corrects for sample overlap that can inflate genetic correlations, improving the accuracy of genetic correlation estimates [19].

Genomic SEM relies on summary-level GWAS data as its primary input along with ancestry-matched LD scores (we used HapMap3 SNPs, [19]). Through the application of LDSC, Genomic SEM identifies genetic covariance patterns within and between all traits. Building on the idea of structural equation modeling, which allows us to systematically analyze relationships among (measured) variables, Genomic SEM translates genetic variance of various traits into a common genetic factor (as depicted in Figure 1). The contributions of the included traits are reflected by their factor loadings. The residual variance captures the genetic signatures unique to specific traits, representing genetic influences that are not shared across all traits. For a detailed explanation of Genomic SEM please see [19]. We extended factor models to include various mediators, categorized as biological (serotonin (SER) [20], cortisol (COR) [21], growth hormone receptor (GHR) [22], C-reactive protein (CRP) [23], interleukin (IL)-6 [24], IL-8 [25], Tumor necrosis factor *α* (TNFA) [26], BDNF [27]), lifestyle and behavioural (alcoholic beverages per week (AI) [28], alcohol dependence (AD) [29], smoking initiation (SI) [28], BMI [30], waist-to-hip ratio (WTH) [31], insomnia (IS) [32]) and psychosocial factors (loneliness (LN) [33], educational attainment (EA) [34]), to characterize underlying correlations (Table 1). The choice of mediators investigated in the current study rests upon prior research e.g. the possibility of physiological changes following insomnia [35], immunosuppressive consequences of stress, as extensively researched in literature (e.g. [36] and [37]), and upon data availability.

**Figure 1:**
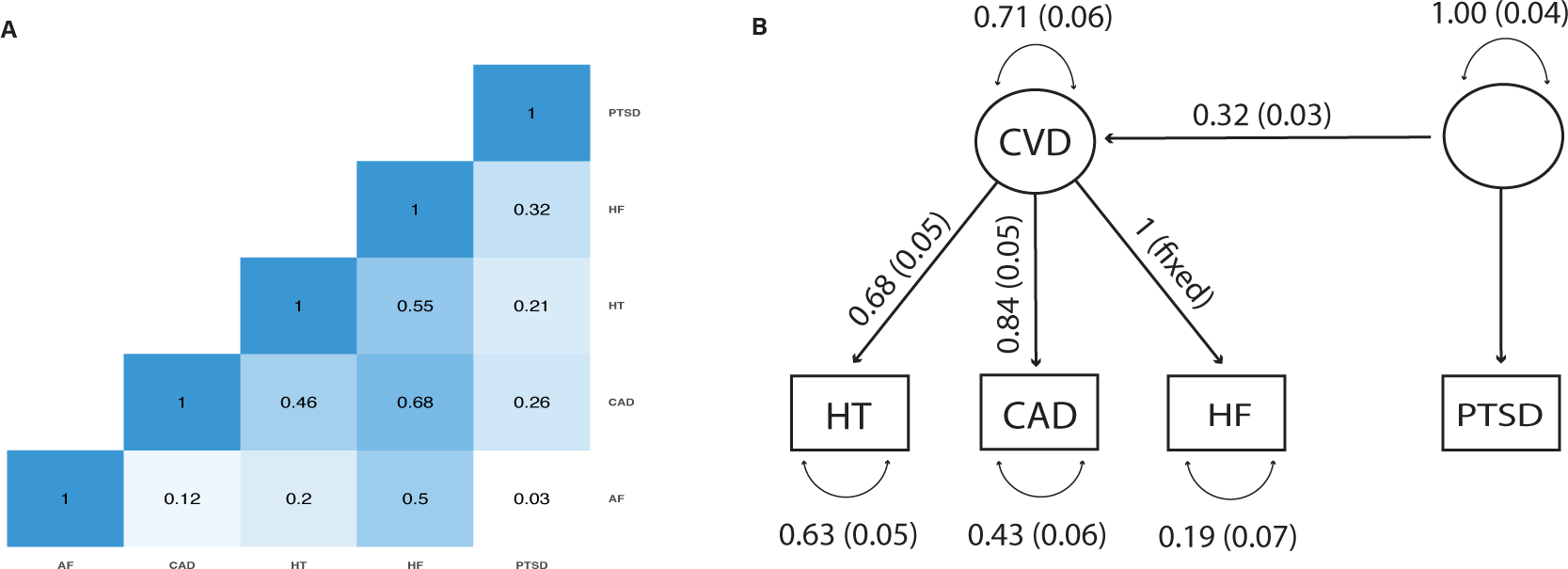
Genomic SEM analyses of cardiovascular traits and PTSD. **A** Genetic correlation of cardiovascular traits, that is, atrial fibrillation (AF), coronary artery disease (CAD), hypertension (HT), and heartfailure (HF), with post-traumatic stress disorder (PTSD). Factor analysis of cardiovascular traits and PTSD (**B**). We found modest genetic correlation of PTSD with HT, CAD, and HF (*r_g_* = 0.21 (p = 3.21 *·* 10*^23^), *r_g_* = 0.26 (p = 2.29 *·* 10*^37^) and *r_g_* = 0.32 (p = 3.08 *·* 10*^16^) respectively, see Figure 1A). Between PTSD and AF, on the other hand, there was no clear evidence for genetic correlation (*r_g_* = 0.03 (p = 0.16), see Figure 1A), which is why we disregarded AF in the following Genomic SEM models. Using Genomic SEM, we fitted a model in which all remaining cardiovascular traits are represented by a shared latent factor. The genetic variance of the individual traits to the common genetic factor were consistently high (factor loadings *>* 0.4).Estimating correlation of the PTSD GWAS on the common cardiovascular factor revealed that 10.2% of the variance in the cardiovascular latent factor are shared SNP effects with PTSD (Figure 1B).

In a follow-up analysis, we aimed to elaborate on direct effects of PTSD on cardiovascular traits within our Genomic SEM framework. To do so, we used the common factor of CVD traits and PTSD and included genome-wide significant (*p <* 5 *·* 10*^8^), independent (*r*^2^ = 0.01) SNPs of the PTSD GWAS through the PTSD indicator itself on the separate cardiovascular traits (as introduced by [19]). This allowed to estimate the effect of SNPs that are robustly correlated to PTSD, later used as instrument variables for MR, on individual cardiovascular traits. This provides some evidence on the potential causality of this relationship [19]. To address the potential influence of horizontal pleiotropy, meaning a direct effect of the SNP in question independent of the proposed exposure PTSD, we included direct loadings of the specific SNP on all three cardiovascular traits for each SNP one by one. For final model estimation, we retained all SNP effects that showed significance (at nominal level *p <* 0.05) and removed any that had lost significance in the full model. The resulting final model thus incorporated significant paths from individual models that retained their significance even when considering all SNPs with associations acting directly on cardiovascular traits rather than through PTSD only.

### Causal associations between PTSD and CVD

Finally, we applied two-sample MR in R (R version 4.2.2 (2022-10-31), ‘TwoSampleMR’ version 0.5.7, [39]), aiming to investigate causality underlying the relationship between PTSD and CVD with more certainty. This approach leverages genetic variants that act as proxies for specific risk factors, mimicking the relationship between these risk factors and the outcome of interest. Because genetic variants are randomly transferred from parents to offspring, MR effectively reduces the impact of confounding and bias, offering a powerful tool for inferring causality in epidemiological research and providing valuable insights into the effects of various risk factors on health outcomes. Causal estimates rely on the use of instrumental variables, meaning SNPs that are strongly linked to the exposure (known as the relevance assumption) [40]. We identify SNPs based on their statistical significance (*p <* 5 *·* 10*^−8^) and clump them using a European LD panel (*r*^2^ = 0.01). MR further assumes exchangeability, prompting that these genetic variants should be interchangeable when assessing their impact on the outcome. Furthermore, it assumes that the genetic variant should only affect the outcome through its influence on the exposure, excluding the possibility of horizontal pleiotropy (extensively reviewed e.g. in [40]).

We calculated causal links using inverse-variance weighted (IVW) estimates. To ensure the robustness and credibility of these estimates, we conducted a series of sensitivity analyses, including weighted median [41] and weighted mode [42], which down-weigh or eliminate outliers and pleiotropic effects, thereby providing alternative estimates to the standard IVW method. To address the potential issue of horizontal pleiotropy, we employed the MR-Egger test. The MR-Egger intercept reflects the extent of horizontal pleiotropy, providing a corrected causal estimate with its slope [43]. Additionally, we utilized Steiger analysis [44] to gauge the influence of instrumental variables on the exposure in comparison to the outcome, thus helping to mitigate concerns regarding reverse causality. Finally, MR-PRESSO [45] was implemented to identify and remove outliers from our analysis, further enhancing the integrity of our results [40]. To avoid sample overlap and consequent infla-tion, we used the PTSD GWAS summary data excluding UKB for all mentioned methods. However, to increase power, we also used the GWAS summary data including UKB while correcting for the overlap by using the method MRlap. MRlap enhances our analysis by adjusting the IVW estimate, compensating for biases due to sample overlap through the deployment of cross-trait LDSC [46].

We define a reliable causal effect if direction and magnitude of the effect are comparable among all (sensitivity) methods. To rule out reverse causal effects of CVD on PTSD we conducted bidirectional MR. We investigated mediating roles of aforementioned factors using multivariable MR (MVMR) [47]. MVMR, an extension of MR, allows for estimating the causal effects of multiple exposures on an outcome by assessing the direct causal effects of each exposure. This allows to assess whether the total effect can partly be explained by certain (risk) factors. To perform MVMR, a set of SNPs associated with the exposure variables but not directly impacting the outcome are utilized to predict the exposures and estimate their effects on the outcome through multivariable regression analysis [47]. We found 46 independent significant SNPs for the PTSD GWAS (excluding UKB) and computed instrument strengths (F) and Cochran’s heterogeneity (Q). We instrumented 37, 36, 29 and 42 variables for the analysis of the effect of PTSD on AF, HF, HT, and CAD respectively. Initially, we could only identify 18 instrumental variables for HT, so we looked for proxies of remaining SNPs, finding an additional 11 instruments (LD *>* 0.9). To examine the possibility of a reverse causal effect we investigated the effect of AF, HF, HT, and CAD on PTSD respectively with 112, 10, 245, and 224 instrumental variables respectively. To ensure robustness and exclude instruments strongly correlated with the outcome, we performed Steiger filtering and repeated parameter estimation. 34, 27, 26 and 33 instrumental variables remained after Steiger filtering for the effect of PTSD on AF, HF, HT, and CAD respectively. Similarly, 112, 10, 245 and 224 instruments remained for the analysis of the effect of CVD on PTSD.

## Results

### Genetic correlation and latent factor analysis

To assess mediating mechanisms linking PTSD to the cardiovascular traits and their common factor, we first examined genetic correlations of PTSD and CVD traits with all mediators. We found low genetic correlation for Serotonine, Cortisol, Growth-hormone receptor, Brain-derived neurotrophic factor, TNF-*α*, alcohol intake and BMI. C-reactive protein, IL6, IL8, alcohol dependence, smoking initiation and insomnia showed moderate genetic correlation and loneliness high genetic correlation with PTSD (with high *< −*0.6 *<* medium *< −*0.2 *<* low *<* 0.2 *<* medium *<* 0.6 *<* high, see Figure S1). We report changes in shared genetic correlation in percentage when accounting for variance covered by the mediator. First, we regarded cardiovascular traits individually (sketched in Figure S2A). We found that shared genetic variation of CAD and PTSD is mainly mediated by behavioural traits, with insomnia (Δ = 30.29%) and educational attainment (Δ = 27.19%) showing the largest effect. The association of PTSD with HF appeared to be mediated mostly by waist-to-hip ratio (Δ = 36.49%), followed by C-reactive protein (Δ = 25.94%), smoking initiation (Δ = 25.83%) and IL6 (Δ = 25.70%). The latter also played a leading mediating role in the correlation with HT (Δ = 39.37%), accompanied by insomnia (Δ = 39.19%) and again waist-to-hip ratio (Δ = 35.89%). Regarding mediators affecting the association of PTSD and the common genetic factor of CVD allows to draw conclusions of PTSD on general CVD rather than on individual traits (depicted in Figure S2B). We found, that insomnia (Δ = 30.37%) and waist-to-hip ratio (Δ = 29.81%) showed most mediation, followed by educational attainment (Δ = 26.43%), IL6 (Δ = 25.55%), and C-reactive protein (Δ = 24.87%). Smoking (Δ = 20.21%) and alcohol dependence (Δ = 17.52%) also play a significant role (Figure 2).

**Figure 2:**
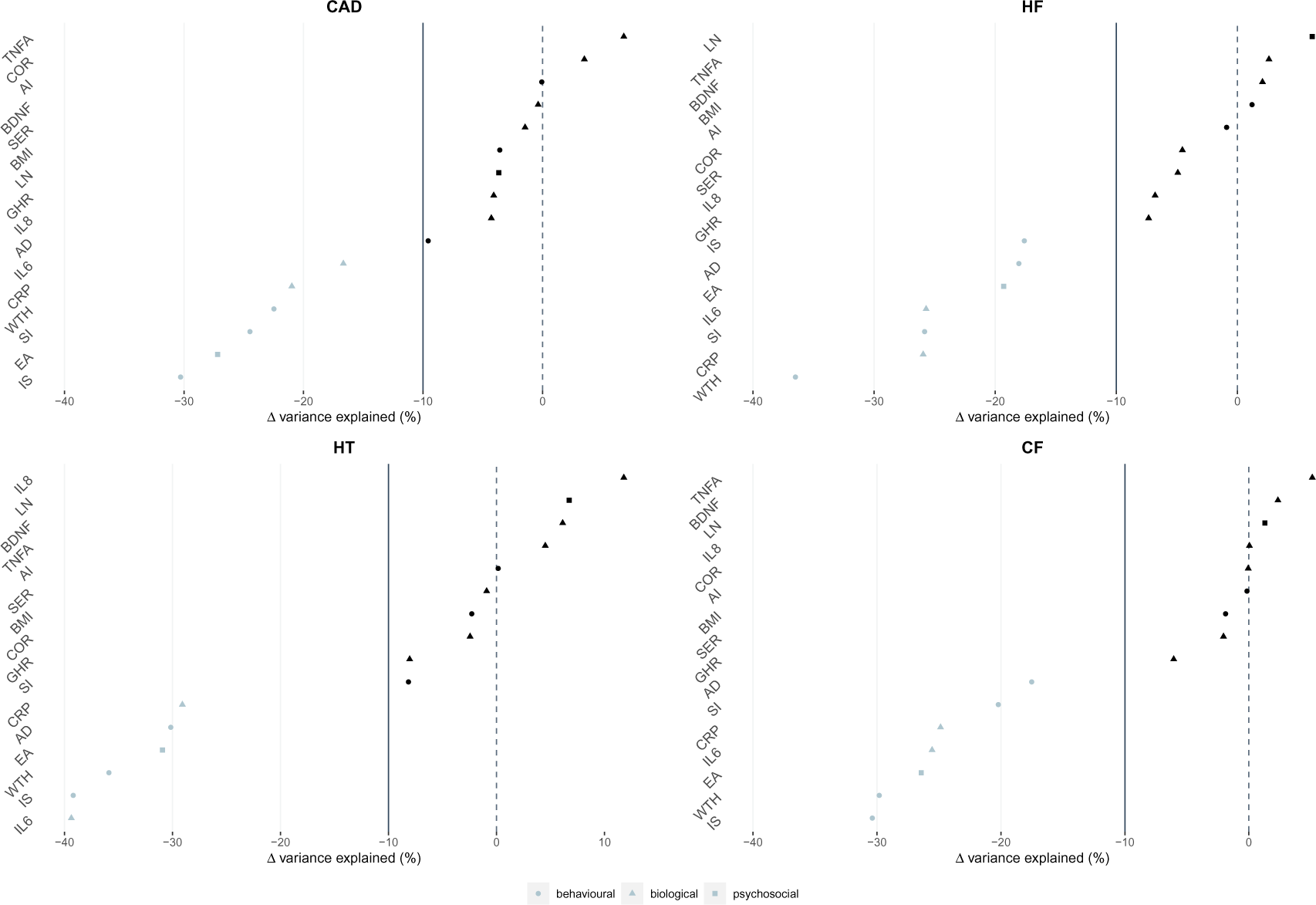
Extension of (factor) models to include various mediators, categorized as biological (serotonin (SER) [20], cortisol (COR) [21], growth hormone receptor (GHR) [22], C-reactive protein (CRP) [23], interleukin (IL)-6 [24], IL-8 [25], Tumor necrosis factor *α* (TNFA) [26], BDNF [27]), lifestyle and behavioural (alcoholic beverages per week (AI) [28], alcohol dependence (AD) [29], smoking initiation (SI) [28], BMI [30], waist-to-hip ratio (WTH) [31], insomnia (IS) [32]) and psychosocial factors (loneliness (LN) [33], educational attainment (EA) [34]), to characterize underlying correlations. We show deviations in the variance, reported as percentage change, shared between PTSD and individual CVD traits when correcting for these mediators (coronary artery disease (CAD), heart failure (HF), hypertension (HT), depicted in Figure S2A). Further, we report mediation of the relationship of PTSD and a common genetic factor of cardiovascular disease (CF) (depicted in Figure S2B)). We define a change in explained variance exceeding 10% as substantial concluding the specific factor to play a mediating role in the regarded association.

### Causal associations between PTSD and CVD

We found strong evidence for a causal effect of PTSD on CAD (IVW OR: 1.53, 95% CI: [1.19, 1.96], p = 0.001) consistent across all (sensitivity) methods (Figure 3, Table S1). The MR-Egger intercept shows no evidence of horizontal pleiotropy (p = 0.90, Table S2). However, the *I*^2^ statistic indicates an unreliable Egger estimate (*I*^2^ = 0.24). Results of the analysis of Steiger filtered instruments confirmed PTSD as a causal risk factor for CAD (IVW OR: 1.37, 95% CI: [1.17, 1.61], p = 0.0001, Figure S3, Table S3). To assess the role of sample overlap in this results, we performed MRlap. The resulting IVW estimate corrected for sample-overlap also indicates a significant causal effect of PTSD on CVD, slightly smaller in magnitude (corrected OR: 1.28, 95% CI: [1.17, 1.40], p = 6.68 *·* 10*^8^).

**Figure 3:**
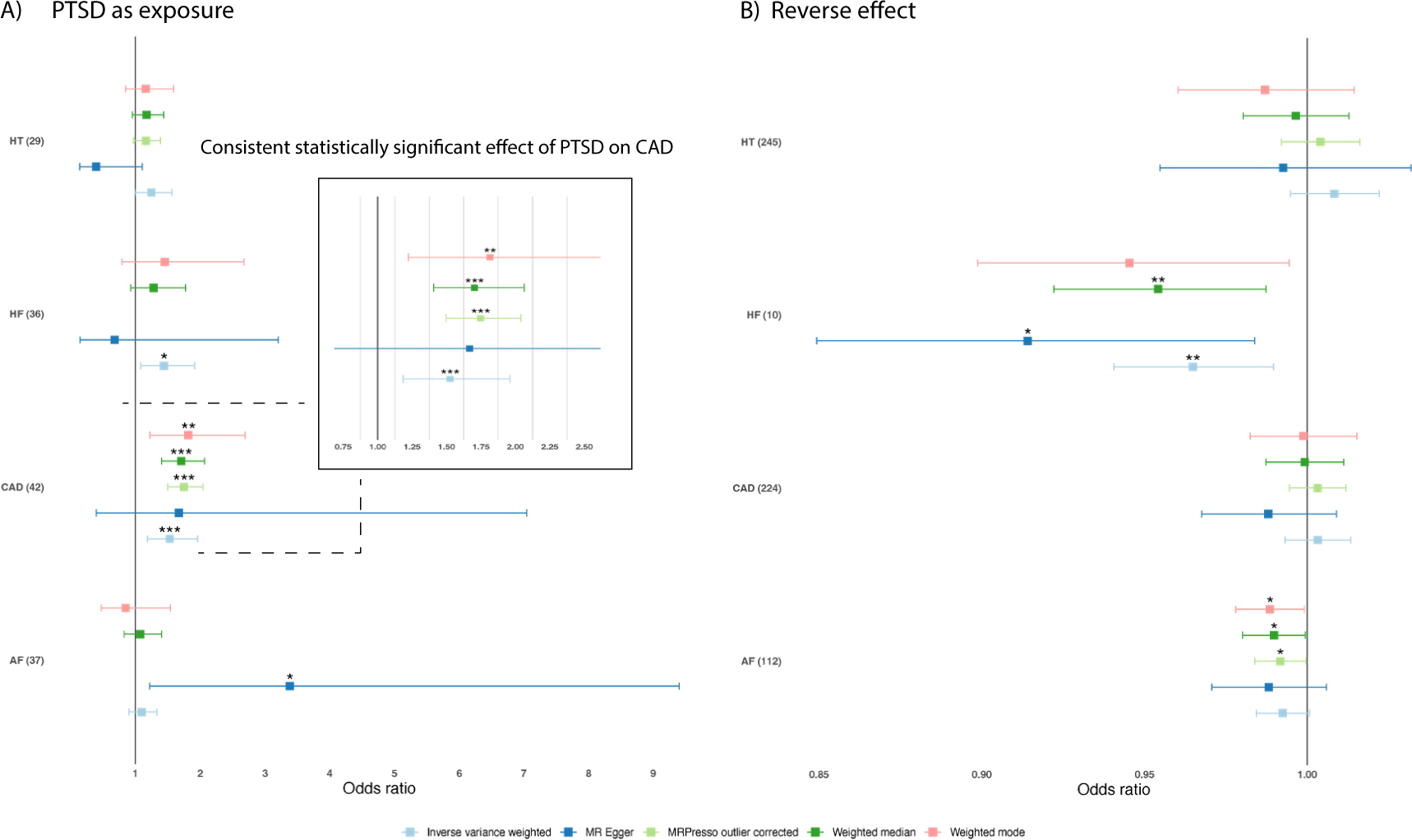
Analysis of Instrumental Variables of MR for the effect of **A** PTSD on various CVD traits and **B** of various CVD traits on PTSD. Number of facilitated instrumental variables in brackets.\**p* < .05, \*\**p* < .01, \*\*\**p* < .001.

Our MR analysis further indicated evidence for causality of PTSD on HT (IVW OR: 1.25, 95% CI: [1.05, 1.49], p = 0.012) (Figure 3) consistent across weighted median, weighted mode, and MR-PRESSO. MRlap supports this finding (corrected OR: 1.27, 95% CI: [1.09, 1.47], p = 0.002). While MR-Egger indicated a point estimate OR *<* 1, this analysis was unreliable with an *I*^2^ statistic of *<* 0.6 (*I*^2^ = 0.25). Analysis of Steiger-filtered data did not produce significant estimates (Figure S3, Table S3). We also found evidence for a causal effect of PTSD on HF (IVW OR: 1.44, 95% CI: [1.08, 1.92], p = 0.012), consistent across IVW, weighted mode, weighted median and MR-PRESSO. While the point estimate of MR-Egger was contrasting in direction we again note unreliability of MR-Egger (*I*^2^ = 0.29) as well as wide confidence intervals. The MR-Egger intercept showed weak evidence of horizontal pleiotropy (p = 0.06, Table S2). Analysis of Steiger-filtered data indicated no evidence of significant effects of PTSD on HF (Figure S3). For both relations, PTSD on CAD and on HF, Cochren’s Q-statistics shows significant heterogeneity for instruments (Table S2). All instrumental variables were of sufficient strength (F-statistics *>* 10; Table S2).

We extent our exploration of causality in the relationship of PTSD and CVD by implementing individual SNP effects into our previously defined common genetic factor model from our Genomic SEM analysis. We included genome-wide significant independent SNPs of the PTSD GWAS on the PTSD indicator in the model. We found significant (*p <* 0.05) loadings on all regarded CVD traits with the largest effect seen on HF followed by CAD. Variance in HT explained by PTSD-specific SNPs is negligibly small. As reverse causality is unlikely when facilitating genetic markers like SNPs, we conclude that these results indicate that a causal cascade from PTSD on CAD and HF may exist. Among the SNPs robustly associated with PTSD, 16 demonstrated significant direct effects on cardiovascular health, thus acting on disease traits through paths distinct from PTSD. Most of these horizontal pleiotropies identified effects on CAD (Figure 4). In other words, these SNPs affect both PTSD and CAD, but the effects on CAD are not simply a consequence of their influence on PTSD. This finding implies that there are distinct biological pathways through which these SNPs exert their effects on CAD, separate from any pathways involving PTSD.

**Figure 4:**
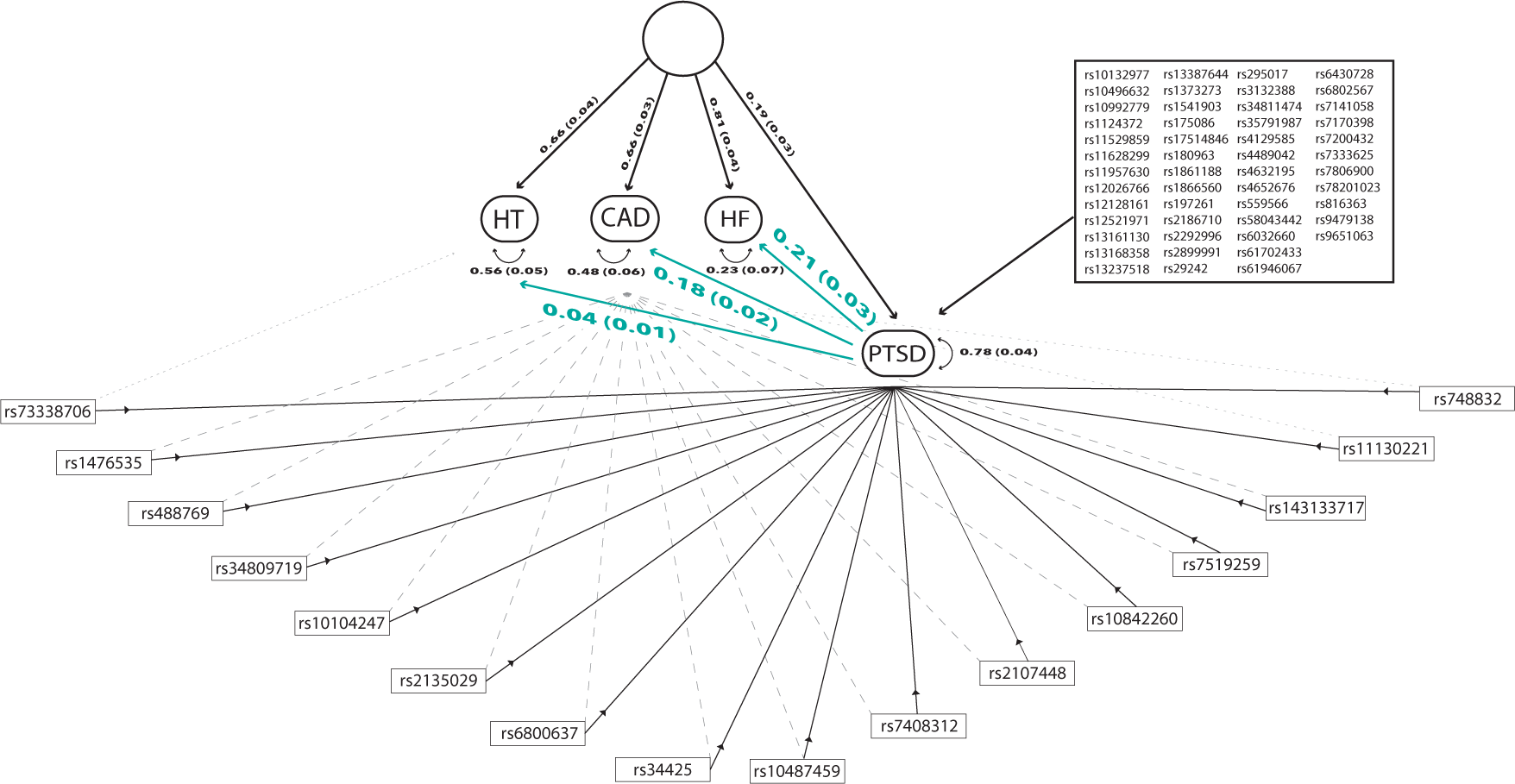
Extension of the common factor model of cardiovascular traits and PTSD by including signif-icant independent SNPs from the PTSD GWAS. Significant associations were found between PTSD and cardiovascular traits, with the most pronounced effects observed on HF and CAD. Furthermore, 16 SNPs were identified that directly influence cardiovascular traits, independently of their effects on PTSD. Green factor loadings ultimately represent directional effects of PTSD-specific SNP effects on individual cardio-vascular traits. All indicated estimations are significant (p *<* 0.05) underscoring the robustness of our results. Variance in HT explained by PTSD is found to be negligibly small. Our findings indicate a causal association between PTSD and both CAD and HF.

MVMR analysis revealed that none of the mediators removed the entire effect of PTSD on CVD. Results indicated that insomnia (Δ = 19.86%) and smoking initiation (Δ = 23.59%) mediate part of the causal effect of PTSD on CAD. For the effect of PTSD on HF we found that smoking initiation (Δ = 42.35%) showed a change in effect, again agreeing with the Genomic SEM mediation models. Multivariable analysis with HT indicated alcohol intake (Δ = 41.09%), insomnia (Δ = 35.79%) and BDNF (Δ = 20.83%) as possible drivers of causality. Mind that changes in effects reported are point estimates, confidence intervals of both the uni- and multivariable effects overlap for each mediator, allowing only soft indications of possible mediation. Mind that sufficient instrument strength was reached for a subset of the analyzed mediators only (F *>* 10 for IL6, AI, IS, SI for CAD, HF and HT and BDNF for HT, Figure S4).

To examine the possibility of reverse causality, we performed MR analysis with CVD as the exposure and PTSD as outcome Table S4. All instruments were of sufficient strength (F-statistic *>* 10; Table S5). We found evidence that HF causally decreases the risk of PTSD (IVW OR: 0.96, 95% CI: [0.94, 0.99], p = 0.006, Figure 3, Table S4), a direction of effect that is unexpected and not in line with previous research. There was no strong evidence for horizontal pleiotropy from the MR Egger. Cochran’s test showed no evidence of heterogeneity (Table S5). *I*^2^ statistics *>* 0.6 implied the need to apply Simex correction, which showed significant p-values associated with the effect on HF. MR on Steiger-filtered instruments, supported the role of HF on PTSD (IVW OR: 0.96, 95% CI: [0.94, 0.99], p = 0.006, Figure S5, Table S6).

## Discussion

This study was undertaken with the overarching goal of establishing the nature of the link between PTSD and CVD. We found notable genetic correlations of PTSD with HT, HF, and CAD, but not between PTSD and AF. Insomnia, waist to-hip ratio, educational attainment, smoking initiation and alcohol dependence along with inflammation indicators like IL6 and C-reactive protein, partially mediated these genetic associations. Using Mendelian Randomization, which allows causal inference, we found strong and consistent evidence for causal effects of PTSD on CAD and HF risk, and somewhat weaker for causal effects of PTSD on HT. With multivariable Mendelian randomization these causal effects too were partly mediated by lifestyle and biological factors, but importantly, independent causal effects of PTSD on CVD also remained.

While AF is more common among people with PTSD (e.g. reviewed in [48]), in our study AF did not exhibit a significant genetic correlation with PTSD. This could be because there is truly no genetic overlap between the two, or because the genome-wide genetic correlation was not able to capture the presence of localized opposing genetic correlations or divergent biological pathways between AF and PTSD. Although AF shares risk factors and biological mechanisms with the other CVD traits [49], its aetiology is predominantly electrophysical, distinguishing it from hypertension, coronary artery disease, and heart failure, which have more multifactorial causes. This distinction could contribute to the observed difference in genetic correlation with AF. As expected, AF did show a substantial genetic correlation with HF, which are known to co-occur. In future studies, advanced analytical techniques such as factor analysis or GWAS-by-subtraction [50] could provide valuable insights into these observed disparities in future investigations.

We found particularly strong evidence that PTSD is a causal risk factor for developing CAD and HF (which can be the result of CAD). Whereas observational associations between PTSD and CVD have been published by others (e.g. [51] and [52]), we now present convincing evidence, using the sophisticated method of MR, that PTSD causally increases CVD risk. Our evidence is strengthened by the fact that we did not find reverse, increasing effects of CVD on PTSD and is further corroborated by consistent results obtained using a novel extension of the innovative Genomic SEM framework. The consistency of findings across varied methodologies reinforces the validity of our conclusion. The knowledge on causality that we put forth is important because it can lead to targeted interventions that address mental health as a significant risk factor in CVD prevention and management. Currently, despite their heightened risk of CVD, mentally ill patients are notably under-served by clinical procedures, receiving substantially poorer screening and treatment [53]. Our study serves as important evidence that clinical intervention can aid to prevent CVD onset in PTSD patients.

Our MR results also showed evidence for a casual effect of HF on PTSD. As previously noted, it is important to consider potential biases in the results due to the fact that the PTSD measure we used includes both DSM-4 and DSM-5 diagnoses. While the DSM-4 included CVD as a potential factor contributing to PTSD, the DSM-5 subsequently revised this classification, specifically excluding PTSD cases due to CVD [1, 11]. The fact that PTSD caused by CVD may have been excluded in clinicians assessment and electronic health records may have induced a negative association from HF to PTSD. All in all, our results at least indicate that the comorbidity between PTSD and CVD is not due to CVD increasing PTSD risk.

In our study, we identified several critical mediators that explain a significant proportion of the (genetic) relationship between PTSD and CVD, offering deeper insights into their interconnectedness. Previous research has consistently shown increased inflammation to be associated with a higher risk of developing CVD [54]. Combined, our Genomic SEM and Mendelian randomization results now indicate that IL6 and CRP play a mediating, role in the causal relationship from PTSD on CVD. Our results further emphasize the role of insomnia as a critical mediator, confirming the known long-term effects of poor sleep on CVD risk. Prior evidence suggested that poor sleep is associated both with PTSD and CVD, possibly manifesting as autonomic nervous system dysregulation [35]. With our evidence of PTSD causally increasing the risk of CAD, we here emphasize an adverse effect of poor sleep quality and accompanying physiological alterations on cardiovascular health.

Another important mediator that we highlighted is fat distribution. Prior research had noted that fat distribution, rather than BMI alone, may play an essential role in the predisposition for various diseases including cardiometabolic disease [31]. We here confirm this finding as we found that waist-to-hip ratio played a significant mediating role in the relationship between PTSD and the overarching cardiovascular factor, whereas BMI did not. The finding that insomnia and waist-to-hip ratio significantly mediate the causal pathway from PTSD to CVD is highly valuable, as it suggest that sleep quality and fat distribution should be focused on in the prevention of CVD in PTSD patients. Finally, our research revealed mediating, causal effects of both smoking initiation and alcohol dependence on the association between PTSD and CVD. Further, we found that educational attainment partially explained the association of PTSD and CVD. This indicates that a higher socioeconomic status may play a protective role against CVD among PTSD patients.

This study shed light on the directionality and mechanisms underlying the association between PTSD and CVD. LDSC, used for studying genetic correlations, effectively identifies genome-wide associations between PTSD and cardiovascular traits, independent of sample overlap. Unfortunately, LDSC requires data stemming from similar ancestry to operate limiting its applicability in trans-ancestry samples [18]. Here, we implemented LDSC on European ancestry GWAS summary statistics only. Extending this, Genomic SEM allows the linking of multiple traits but inherits LDSC’s limitations and demands large sample sizes and intensive computation citepgrotzinger2019genomic. Neither method accounts for epigenetic or environmental factors in disease correlation. We further applied MR, which provides stronger evidence for causal effects than observational studies, making it invaluable for policy and clinical guidelines [40].

In summary, our findings showed shared genetic liability between PTSD and CVD, as well as strong evidence for causal effects of PTSD on CVD, for CAD and HF in particular. We identified biological and behavioural mediators of the causal cascade of PTSD on CVD which holds significant value for future clinical implications. We highlighted that monitoring waist-to-hip ratio, inflammatory markers, smoking, alcohol use, and sleep quality is essential for early intervention and prevention of CVD in these patients. Ultimately, this study serves as a proof of an increased vulnerability of PTSD patients for CVD and calls to act accordingly in the clinical setting.

## Data Availability

Facilitated GWAS summary statistics were obtained from online resources or through respective GWAS authors

## Acknowledgements

The authors thank the PGC-PTSD investigators for making their data available. Major financial support for the PGC-PTSD workgroup was provided by the Cohen Veterans Bioscience, Stanley Center for Psychiatric Research at the Broad Institute, and the National Institute of Mental Health (NIMH; R01MH106595, R01MH124847, R01MH124851). The authors further thank Michel G Nivard (Department of Biological Psychology and EMGP+ Institute for Health and Care Research, Vrije Universiteit Amsterdam, Amsterdam, the Netherlands) for valuable feedback.

## Data availability statement

Where available publicly available GWAS data was used. In absence of downloadable summary statistics, we contacted corresponding authors. PTSD GWAS summary statistics will be available on the website of the Psychiatric Genomics Consortium (https://pgc.unc.edu/for-researchers/download-results/).

## S1 Supplementary notes

### Data preparation and analysis

Included traits and their respective GWAS are summarized in Table 1. Slight alterations to the preregistered data utilization were necessary as no effect size was published for cy-tokine meausurements [55] and one dataset contained mainly rare variants for mediacation (non-) adherence [56]. These datasets were excluded. A small deviation from the published preregistration included the usage of a larger dataset for alcohol intake [28]. For MR analysis we facilitate both alcohol intake and smoking initiation from the same authors excluding UK Biobank entries to avoid sample overlap with UKB-containing cardio traits in multivariate MR. As the largest and newest GWAS available of educational attainment contains UKB [34], we facilitated their earlier disocvery sample for MVMR to avoid sample overlap [38]. Data preparation for MR included filtering for *p <* 5 *·* 10*^−8^ and subsequent clumping (*r*^2^ = 0.01) using the R-package TwoSampleMR [39].

**Figure S1:**
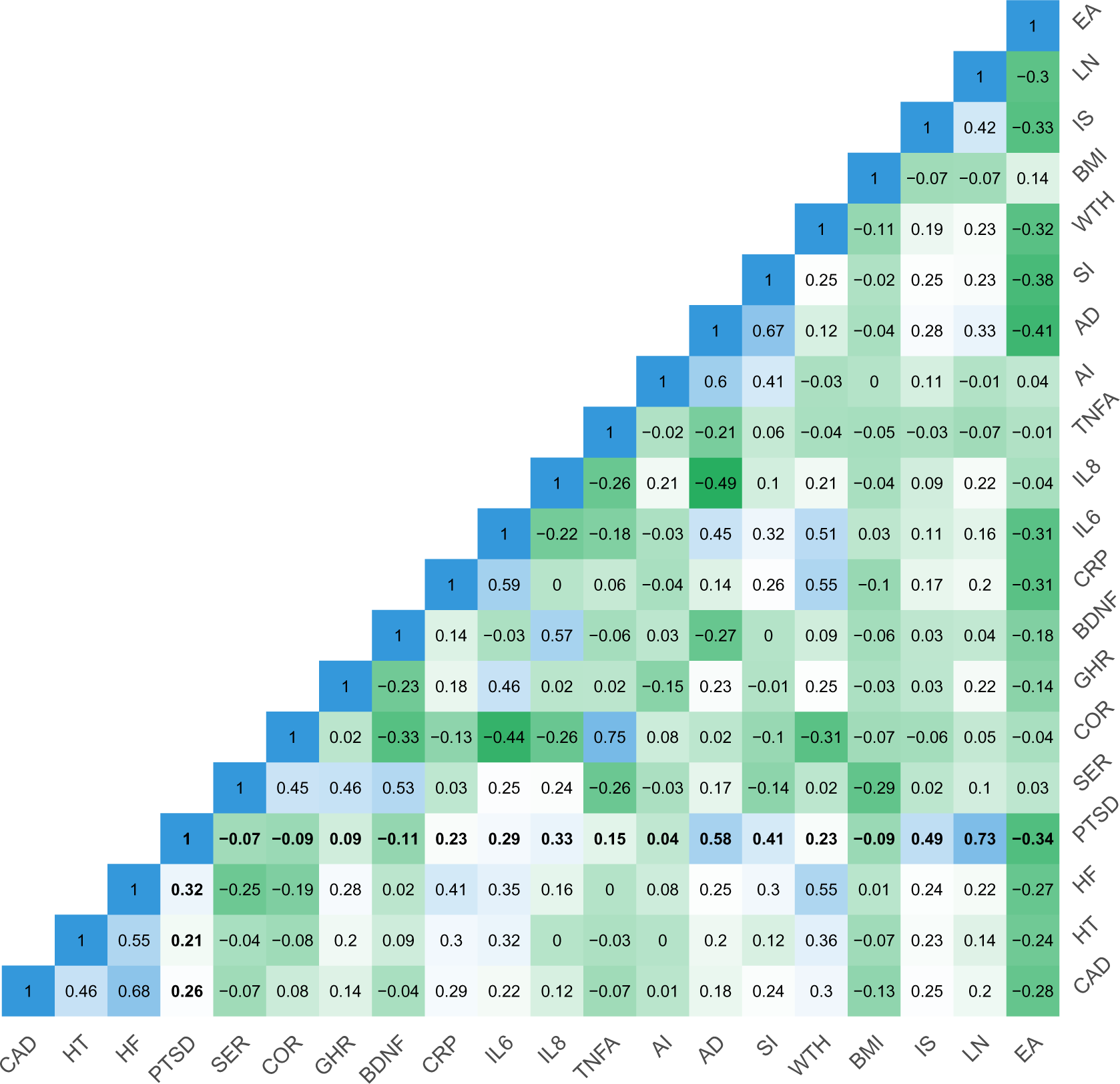
Genetic correlation performed as LDSC for all relevant traits sed for Genomic SEM analysis.

**Figure S2:**
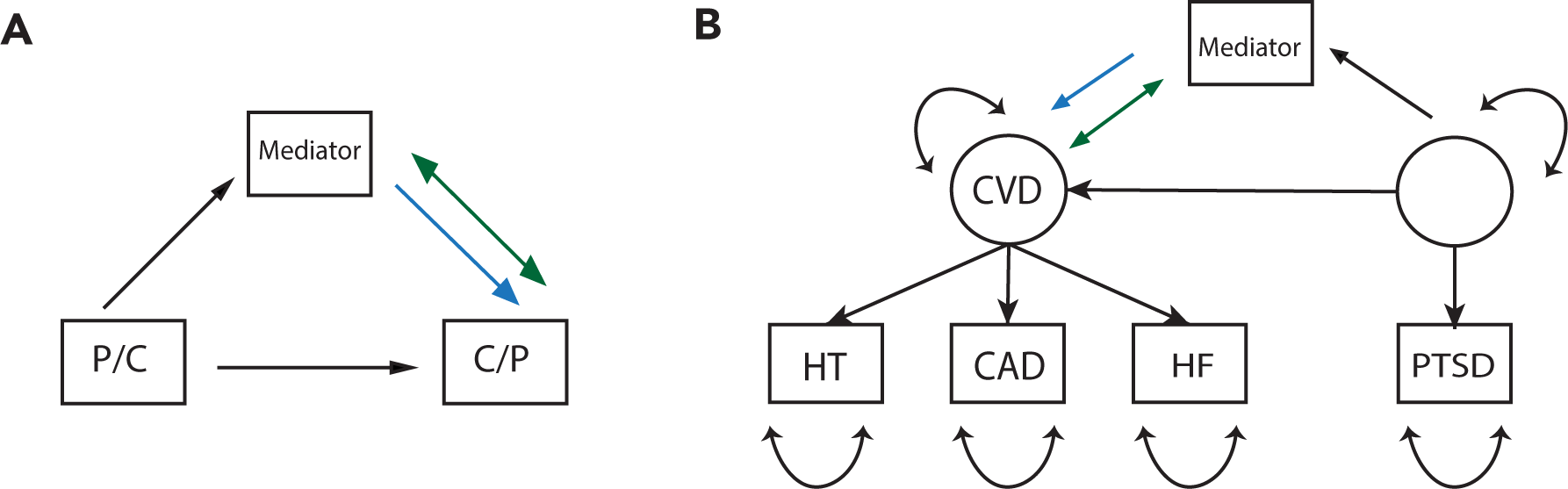
Correlation (green) and mediation (blue) models for **A** individual cardiovascular traits and PTSD, for **B** a common cardiovasular factor and PTSD and for **C** a common cardiovascular and PTSD factor.

**Figure S3:**
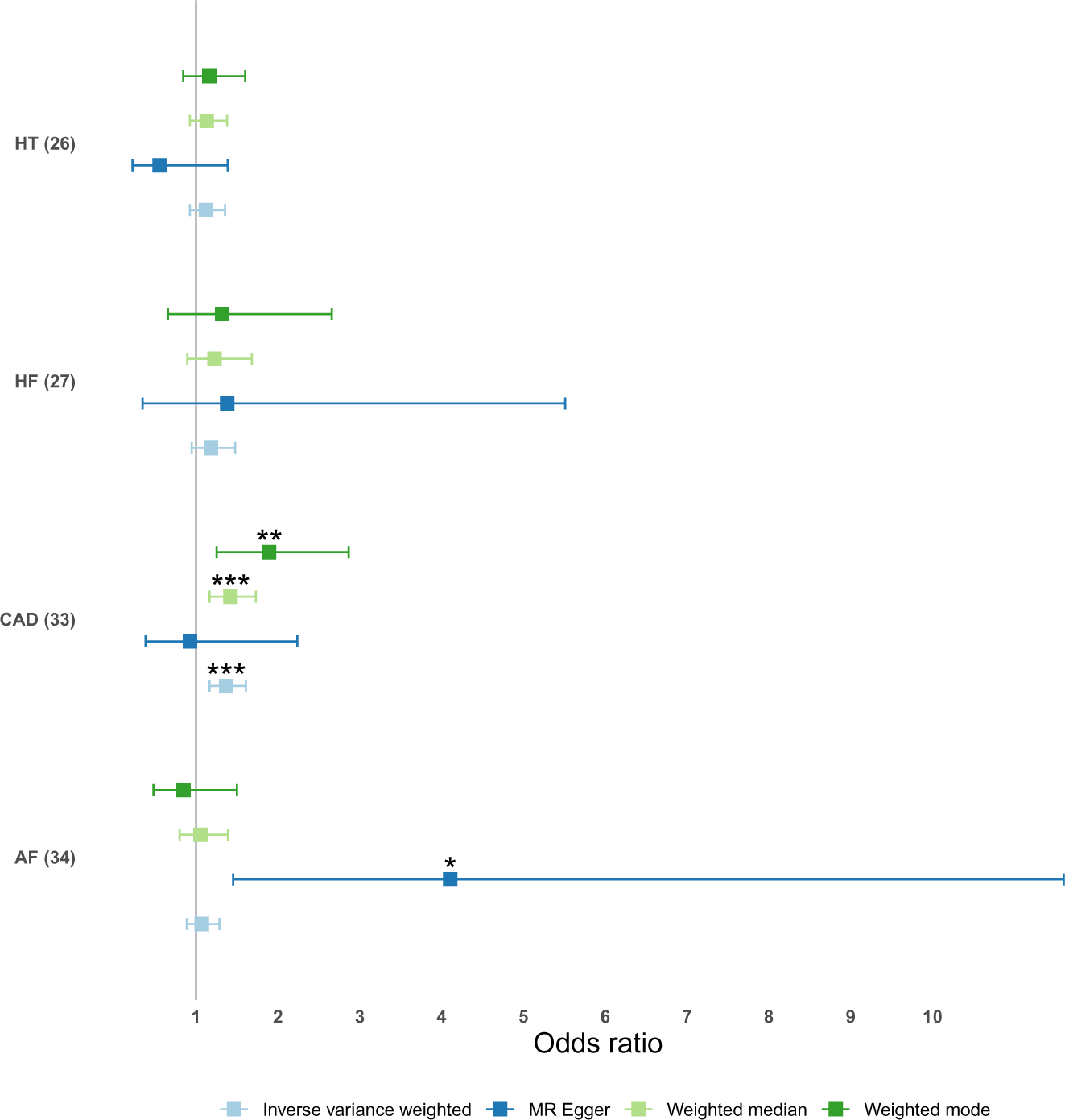
Analysis of Steiger filtered instrumental variables for the effect of PTSD on various CVD traits (number of facilitated SNPs). \**p* < .05, \*\**p* < .01, \*\*\**p* < .001.

**Figure S4:**
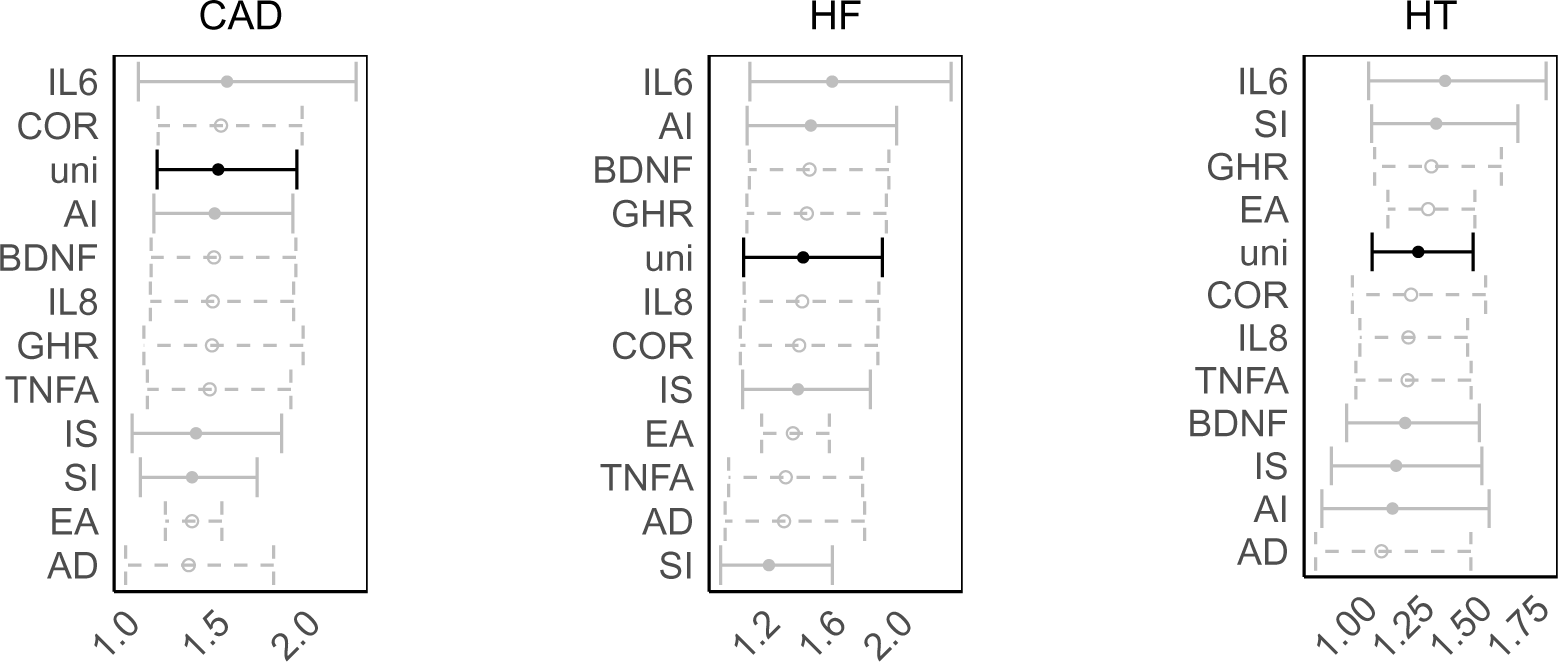
Multivariate MR analysis of the effect of PTSD on CVD traits that previously showed a significant IVW. Dashed lines indicate insufficient instrument strengths measured by F-statistics falling below 10.

**Figure S5:**
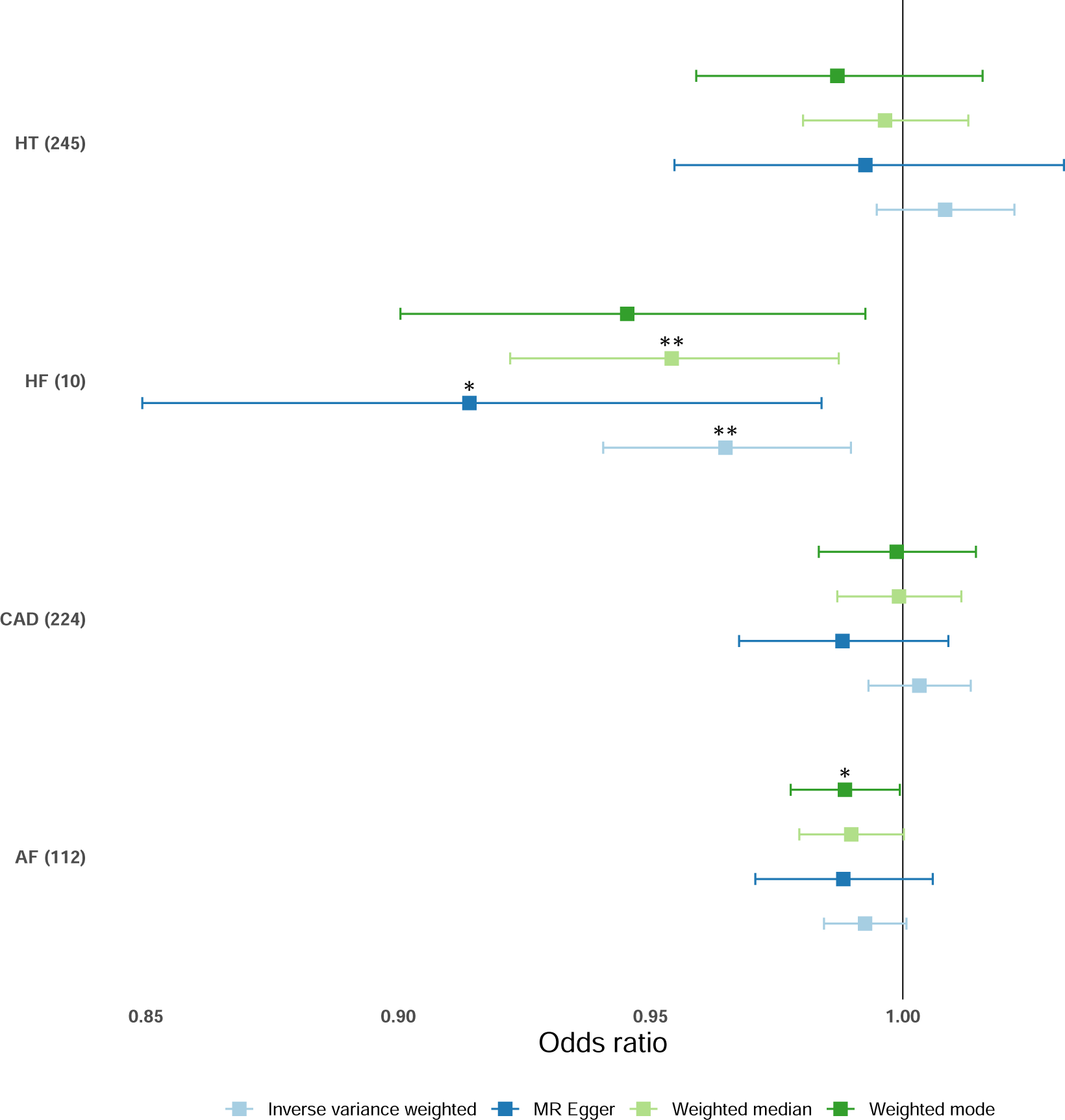
Analysis of Steiger filtered instrumental variables for the effect of various CVD traits (number of facilitated SNPs) on PTSD. \**p* < .05, \*\**p* < .01, \*\*\**p* < .001.

